# Challenges and Facilitators for Physicians and Medical Schools to Promote Social Accountability in Rural Communities: A Scoping Review and Thematic Analysis

**DOI:** 10.1101/2025.01.04.25319991

**Authors:** Aghna Wasim, Ali Abud, Yihan Wang, Ehsan Tavakoli, Athena Moreno-Gris, Samar Joshi, Arthur Wang, Angela Neasadurai, Nikki Shaw

## Abstract

**Background:** Social accountability aligns the health demands and priorities of the community and the commitment of medical schools. Existing literature has proposed contextually distinctive interventions encompassing geographical initiatives, acting networks or parties, and systematic categorizations for the implementation of social accountability in a rural setting. Research concerning social accountability implementation strategies has been insufficient. This study provides an overview of the existing literature surrounding the social accountability practice of physicians and medical schools, in rural settings, through evidence of the barriers and facilitators inherent to the rural communities.

**Methods:** This scoping review was conducted following the Arksey and O’Malley Framework. Web of Science, MEDLINE, EMBASE, SCOPUS, and CINAHL were searched for peer-reviewed studies published since 2000.

**Results:** A total of 2698 abstracts were identified, 180 full-text articles were reviewed, and 53 articles met the eligibility criteria across 15 different countries. Strategies used and problems inherent in promoting social accountability across physician practice and medical education are reported.

**Conclusions:** This scoping review synthesizes existing evidence on the barriers and facilitators of social accountability practices in rural settings globally. The identified literature captures recurring themes of medical infrastructure inadequacy, community immersion curriculum design, targeted admission, geographical isolation, and institutional or peer support.

## Introduction

About 45% of the world’s population, amounting to over 3 billion people, live in rural and remote areas where they experience a number of challenges in accessing quality healthcare.^1–4^ These include a lack of proper healthcare facilities, geographical inaccessibility of clinics and hospitals, poor infrastructure, transportation difficulties, and financial constraints.^5–8^ In addition, a majority of the people residing in these areas belong to vulnerable and socioeconomically disadvantaged groups.^9^ Rural communities have generally poorer health compared to urban communities.^9,10^ Individuals are more likely to experience chronic diseases and are less likely to have medical insurance.^11–13^ Similarly, rural populations have high rates of smoking, heavy drinking and obesity.^14–16^ Moreover, rurality has found to be associated with shorter life expectancy.^17^ For years, rural and remote populations have also struggled to recruit and retain physicians, leading to a shortage of healthcare professionals from primary care physicians to specialists and in turn exacerbating existing disparities in health status and access.^11,18^

In recent years, there has been an emphasis on addressing the lack of physicians in rural and remote areas through promoting social accountability in medical education. Social accountability is the “obligation [of medical schools] to direct their education, research and service activities towards addressing the priority health concerns of the community, the region, and/or nation they have a mandate to serve.”^19^ It is a paradigm shift that requires stakeholders such as physicians and medical educators to reassess their personal and collective responsibilities in addition to implementing interventions to reduce the maldistribution of physicians across rural and urban areas and meet the needs of rural populations.^20^ Efforts to incorporate social accountability into medical education have focused on recruiting students that are representative of the local population, teaching social accountability-related material in lectures, increasing community engagement, offering service-learning placements within rural settings, and providing opportunities for international electives.^21^

The importance of social accountability in facilitating equitable access to medical care, empowering marginalized communities and vulnerable groups, and improving public health and quality of care is beginning to receive recognition. Although several studies have examined initiatives to promote social accountability in physicians and medical schools, there have been no attempts to scope and synthesize literature, recognizing factors that might ease or hinder their implementation. This scoping review was conducted with the aim to provide an overview of the literature on the promotion of social accountability in rural and remote areas, specifically identifying challenges and facilitators encountered by physicians and medical schools in the process.

## Methods

### Framework

This scoping review was conducted to identify barriers and facilitators faced by physicians and medical schools while attempting to promote social accountability within rural communities. Scoping reviews are a type of evidence synthesis that provide a comprehensive landscape of the body of literature on a particular topic.^22^ The aim was to provide a broad overview and identify any knowledge gaps, pertaining well to this specific methodology. This review followed the Arksey and O’Malley framework^23^ which consists of six steps: 1) identifying the research question; 2) searching the literature for relevant studies; 3) screening studies for eligibility; 4) data extraction; 5) data analysis; and 6) an optional stakeholder consultation. A stakeholder consultation was not performed.

### Search strategy and study selection

A comprehensive search strategy using keywords such as “social accountability”, “physicians” and “medical school” was tailored to each database including Web of Science, MEDLINE, CINAHL, Scopus and Embase (Supplemental File 1). Title and abstract and full-text screening was conducted on Covidence.^24^ For both stages of screening, each record was independently assessed for eligibility using the predetermined inclusion criteria by at least two reviewers and any conflicts were resolved by consensus. Primary articles of any study design that addressed social accountability among physicians and medical schools in the context of rural and remote communities were included. Articles were only included if they were written in English. On the other hand, articles that did not discuss social accountability from the perspective of physicians or medical schools were excluded. Studies were also removed if they were based in non-rural settings. Moreover, non-research articles (e.g., errata, commentaries, book chapters, letters to the editor, protocols, editorials, perspectives and opinion pieces), thesis dissertations, conference proceedings, and reviews were excluded.

### Data extraction and analysis

A standardized data extraction template was created on Covidence. The extraction form for each study was independently populated by two reviewers with information on the methodological and participant characteristics along with the main findings. A third reviewer collated the abstracted data through consensus. Study characteristics extracted included 1) the title of the article; 2) name of the lead author; 3) the country in which the study was conducted; 4) date of print publication; 5) study design (e.g., cross-sectional, qualitative, mixed-methods, case report etc.); 6) setting (e.g., rural village, medical institution); 7) participant population (e.g., medical educators, physicians, medical students); 8) population size; 9) participant demographics and age; and 10) study objectives. Furthermore, the following information pertaining to social accountability was abstracted from each study: 1) its definition; 2) importance (e.g., impact on patient outcomes or community wellbeing); 3) measures (e.g., interviews, perspectives, surveys etc.); 4) social accountability challenges experienced by physicians and medical schools; 5) the impact of these barriers on social accountability; 6) proposed solutions; 7) key roles to be taken up by physicians, medical schools and educators; 7) facilitators of social accountability for physicians and medical schools; 8) impact of the intervention; and 9) translation of social accountability in other communities (e.g., similar medical education, training frameworks, physician workshops). Information regarding the article’s conclusion, authors’ suggestions, directions for future research and study limitations were also reported in the extraction form. The data was subsequently analyzed using a thematic analysis approach and the findings were reported narratively.

## Results

### Search results

The database search was conducted on April 1st, 2024. The database search yielded 2698 hits from which 692 records were removed due to duplication. The remaining 2006 records were screened for eligibility based on their title and abstracts and another 1826 articles were excluded. 180 full-text records were assessed against the inclusion criteria and a total of 53 articles were deemed eligible for inclusion in the final scoping review (Figure 1).

**Figure 1.**
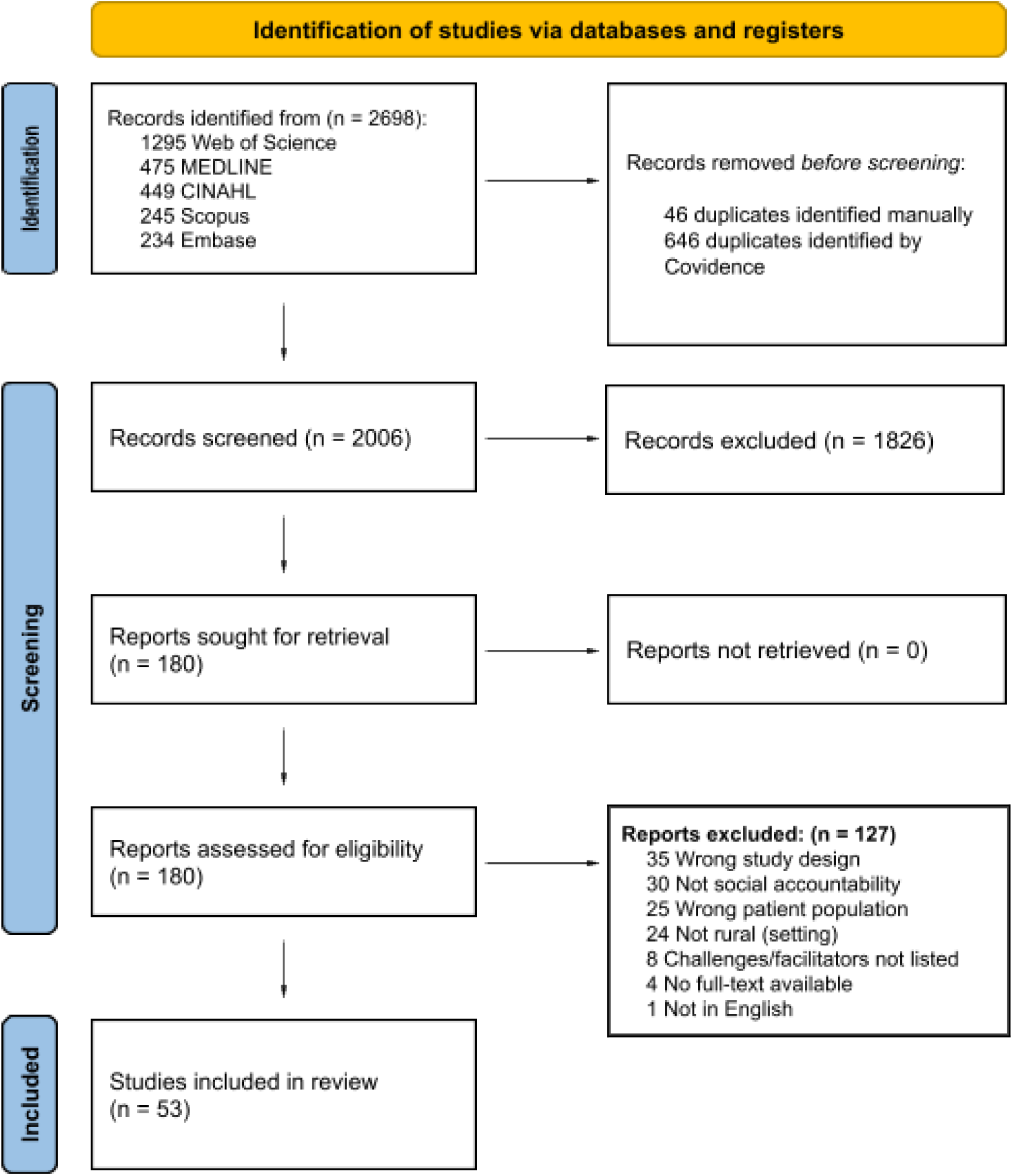
PRISMA (Preferred Reporting Items for Systematic Reviews and Meta-Analyses) Flow Diagram

### Descriptive Summary

All articles were published after 2005 with six (11.3%) articles being published between 2005-2010, 35 (66.0%) articles between 2011-2020 and 12 (22.7%) articles between 2021-2024 (Table 1). A majority (45.3%) of the included articles were qualitative studies. 13 (24.6%) studies followed a cross-sectional design, eight (15.1%) articles used prospective cohorts while four (7.5%) studies employed a mixed-methods approach. Descriptive analysis was used in two (3.7%) studies while retrospective cohort and retrospective case study approaches were taken in one (1.9%) study each (Table 2).

**Table 1.** Original articles (n=53) included in the scoping review.

| Author | Title | Year of Print Publication | Study Aim(s) | Study Design | Country | Key Challenges | Key Facilitators |
| --- | --- | --- | --- | --- | --- | --- | --- |
| Cheek et al [46] | Building a medical workforce in Tasmania: A profile of medical student intake | 2019 | Compared a medical program’s admission process with a social accountability criteria and investigates the implications of widening participation and widening access | Descriptive analysis | Australia | Medical program fails to target applicants outside the designated rural area | Rural placement opportunities, expanded selection criteria |
| Chen et al [47] | The redistribution of graduate medical education positions in 2005 failed to boost primary care or rural training | 2013 | Aimed to determine the effects of the 1997 graduate medical education cap and 2005 redistribution on medical education positions | Descriptive analysis | United States | Redistribution Of graduate medical education positions did not meet rural workforce needs |  |
| Cleland et al [64] | A qualitative study of social accountability translation: From mission to living it | 2024 | Explored perceptions of socially accountable medical school practices | Qualitative | India |  | Communal living to promote community engagement, formal training aligned with community needs |
| Cleland et al [25] | Attracting healthcare professionals to remote and rural medicine: Learning from doctors in training in the north of Scotland | 2012 | Utilized the social cognitive career theory to note the experiences of physician training through the Foundation Programme | Qualitative | Scotland | Isolated rural placements are viewed unfavorably, attitudes like perceived workload and expenses are unappealing | Providing trainees with a sense of responsibility but also reassurance of available aids and supervision |
| Clithero-Eridon et al [48] | Contribution of the Nelson R. Mandela School of Medicine to a socially accountable health workforce | 2019 | Explored the social accountability of 'fit-for-purpose' medical graduates | Cross-sectional | South Africa | Specialization and skills development are indicators of one's willingness to stay | Recruitment of physicians with similar demographic characteristics to the community, leadership training in medical school curriculum |
| Clithero-Eridon et al [60] | Future medical student practice intentions: The South Africa experience | 2020 | Examined the career profiles and background of Nelson R. Mandela medical students | Cross-sectional | South Africa |  | Family medicine emphasis in the curriculum, fostering a sense of attachment to the community, community immersion |
| Crandall et al [26] | Patients: A longitudinal comparison of problem-based and traditional medical curricula | 2007 | Explored the effects of gender and learning environment towards attitude factors such as willingness to practice in underserved areas | Prospective cohort | United States | Progressively negative attitudes | Promote professionalism through opportunities with vulnerable patients |
| Cristobal & Worley [49] | Can medical education in poor rural areas be cost-effective and sustainable: The case of the Ateneo de Zamboanga University School of Medicine | 2012 | Investigated the sustainability and medical workforce of a low-resource medical school with volunteer faculty | Retrospective case study | Philippines | Funding issues, unstable regional conflict | Recruiting locally and implementing a problem-based curriculum in coordination with community members |
| de Leeuw et al [27] | Educating medical students' "hearts and minds": A humanities-informed cultural immersion program in Indigenous experiential community learning | 2021 | How participation in a cultural immersion program influenced students' attitudes towards rural First Nations communities | Qualitative | Canada | Persistent fear of the unknown, lack of familiarity with the local culture | Experiential and immersive experiences to learn from local community leaders |
| Eggleton et al [50] | Development of a rural strategy for an urban-based medical program: A pragmatic reality | 2024 | Determine how urban-based institutions can improve the rural workforce | Qualitative | New Zealand | Poor understanding of rural needs | Rural-oriented education programs |
| Florence et al [51] | Rural health professions education at East Tennessee State University: Survey of graduates from the first decade of the community partnership program | 2007 | Determine the practice and career intentions and attitudes of The Community Partnerships Program | Cross-sectional | United States | Require a great deal of effort and resources | Community partnership program with service-based learning |
| Gladman et al [28] | Measuring organisational-level Aboriginal cultural climate to tailor cultural safety strategies | 2015 | Determine how to promote cultural safety. | Cross-sectional | Australia | A lack of information on the differences between Aboriginal and non-Aboriginal groups | Promoting culturally safe care |
| Green-Thompson et al [61] | The social accountability of doctors: A relationship based framework for understanding emergent community concepts of caring | 2017 | Examined the perceptions of community members on physician's social accountability | Qualitative | South Africa |  | Sufficient training for students, building knowledge and understanding of the people served |
| Griffin et al [29] | Who do you think you are? Medical student socioeconomic status and intention to work in underserved areas | 2017 | Investigated the connection between medical student backgrounds and practice intentions | Prospective cohort | Australia | Very low SES students were less likely to desire to work in underserved regions, student attitudes in relation to family social factors | Curriculum changes to alter students subjective perspective with regard to their willingness to serve vulnerable communities |
| Guignona et al [52] | A curriculum for achieving universal health care: A case study of Ateneo de Zamboanga University School of Medicine | 2021 | Determined the impact of Ateneo de Zamboanga University School of Medicine students on local communities and determined factors linked to social accountability | Qualitative | Philippines | Resistance to curriculum changes from governing bodies and medical schools | Faculty commitment to a community-centred vision, experiential learning curriculum, student engagement in local workforce |
| Halili et al [53] | Addressing health workforce inequities in the Mindanao regions of the Philippines: Tracer study of graduates from a socially-accountable, community-engaged medical school and graduates from a conventional medical school | 2017 | Determined community outcomes of graduates of the socially accountable Ateneo de Zamboanga University School of Medicine and a Philippines medical school | Cross-sectional | Philippines | No local medical school and lack of infrastructure | Medical school selection of local individuals with lower SES status |
| Hay et al [54] | Selecting for a sustainable workforce to meet the future healthcare needs of rural communities in Australia | 2017 | Described the link between intent to practice, selection scores, and student place of origin | Prospective cohort | Australia | Training of individuals unmotivated to serve in rural areas | Selection reforms of applicants of rural origin and incorporating rural involvement during training |
| Hedrick & McHenry-Sorber [30] | Negotiating the Gemeinschaft/Gesellschaft dichotomy: Appalachian medical student perceptions of practice | 2023 | Explored the factors implicating intent to practice for Appalachian medical students | Qualitative | United States | Prevailing sentiments of rural care, limitations of rural practice such as a lack of amenities, finances | Personal connection during rural rotation, faculty role models to influence desirability of rural practice |
| Heller et al [31] | Exploring medical students' perceptions of family medicine in Kyrgyzstan: A mixed method study | 2023 | Explored influences of medical student specialty choice and perceptions on family medicine | Mixed methods | Kyrgyzstan | Attitudes towards Family Medicine specialty | Parents practicing Family Medicine, developing a sense of responsibility and understanding of local healthcare needs |
| Hogenbirk et al [62] | Milestones on the social | 2016 | Evaluated how exposure to | Cross-sectional | Canada |  | Rural Canadian background was |
|  | accountability journey: Family medicine practice locations of Northern Ontario School of Medicine graduates |  | NOSM University’s medical program affected practice location |  |  |  | indicative of rural practice location, Programs like NOSM University’s Postgraduate education forecasted rural practice |
| Hulme-Chambers et al [32] | Medical termination of pregnancy service delivery in the context of decentralization : Social and structural influences | 2018 | Assessed barriers and facilitators of a decentralization initiative with the aim of improving accessibility to pregnancy medical termination services in rural areas | Qualitative | Australia | Concerns regarding quality of care that can be provided with barriers such as wait times and accessibility | Supportive colleagues, partnering with rural-based organizations |
| Johnston et al [33] | Training a fit-for-purpose rural health workforce for low- and middle-income countries (LMICs): How do drivers and enablers of rural practice intention differ between learners from | 2020 | Examined how learner characteristics and country-specific contextual factors are linked with intention to practice in rural areas, practice field, emigration intention, and how these varies by a country’s income status | Prospective cohort | Philippines , Sudan, South Africa and Nepal | Having an international background | Rural background students, underserved lower SES students |
|  | LMICs and high income countries? |  |  |  |  |  |  |
| Larkins et al [65] | Impact of selection strategies on representation of underserved populations and intention to practise: International findings | 2015 | Described how medical school selection strategies impacted underserved community representation and student practice intentions | Prospective cohort | Australia, South Africa, Sudan, Belgium and Philippines |  | Selection strategies beyond academics and more closely aligned with the local population |
| Larkins et al [34] | Practice intentions at entry to and exit from medical schools aspiring to social accountability: Findings from the Training for Health Equity Network Graduate Outcome Study | 2018 | Identified characteristics linked with intention to work with underserved communities for students at Training for Health Equity Network medical schools | Prospective cohort | Australia, Philippines, Sudan, Belgium, Canada and South Africa | High income and urban background students, creating an aligned workforce, Cost dissuades more representative candidates | Local rural background students with lower parental income |
| Maar et al [35] | Enhancing Indigenous health research capacity in northern | 2018 | Described the successes and challenges in the partnership between NOSM's | Qualitative | Canada | Past negative collaborative experiences among local Indigenous community | Building long-term relationships with local communities, Utilizing the Community |
|  | Ontario through distributed community engaged medical education at NOSM: A qualitative evaluation of the community engagement through research pilot program |  | medical education collaboration and Indigenous community sites |  |  | members, Research Ethics Boards time and resource constraints | Engagement Through Research program |
| Maclaren et al [36] | Understanding recruitment and retention of doctors in rural Scotland: Stakeholder perspectives | 2022 | Considered the perspective of stakeholders on issues within Scottish healthcare | Qualitative | Scotland | Family Preferences, Recruitment and retention difficulties | Rural exposure during childhood, Supportive tight-knit community, Family preferences, Rural placements |
| Malau-Aduli et al [63] | Understanding medical students' transformative experiences of early pre-clinical international rural placement over a 20-year period | 2022 | Utilized Transformational Learning Theory as a framework to describe the influence of international rural placement on participants' development and learning | Qualitative | Australia |  | Immersive international rural placements |
| Mathews et al [37] | Work locations in 2014 of medical graduates of Memorial University of Newfoundland: A cross-sectional study | 2015 | Identified the predictors of practice location for 2014 Memorial University medical graduates | Cross-sectional | Canada | Small community's inability to support diverse medical specialties | Rural background, local residency training |
| Matsumoto et al [66] | Policy implications of a financial incentive programme to retain a physician workforce in underserved Japanese rural areas | 2010 | Described how a rural-oriented financial incentive approach affected medical student practice location | Retrospective cohort | Japan |  | Rural service exposure |
| McGrail & O'Sullivan [38] | Increasing doctors working in specific rural regions through selection from and training in the same region: National evidence from Australia | 2021 | Investigated any correlation between entering medicine and/or training in a rural region and physicians current working area, along with the influence of rural training duration | Cross-sectional | Australia | Mismatch of skills, retention issues, enrollment costs, barriers in maintaining high quality rural training related in finances, sufficient infrastructure and working conditions, community engagement, and sufficiently experienced rural supervisors | Rural background students, General Practitioner specialty training |
| McGrail et al [55] | Rural training pathways: The return rate of doctors to work in the same region as their basic medical training | 2018 | Determined how rural training impacts student practice location and the effects of longer rural exposure and having both training and schooling in the same remote area | Prospective cohort | Australia | Selection being unrelated to student's secondary school location | Preferential selection of rurally-based students, longer duration for local rural exposure |
| Meili et al [56] | Teaching social accountability by making the links: Qualitative evaluation of student experiences in a service-learning project | 2011 | Investigated student experiences during the Making The Links program | Qualitative | Canada | A lack of meaningful clinical experience as a result of short rural training | Exposure to rural and low-income communities through making the links program |
| Mian et al [39] | How underserved rural communities approach physician recruitment: Changes following the opening of a socially accountable medical school in northern Ontario | 2017 | Analyzed NOSM's impact on family physician recruitment in rural and underserved areas | Qualitative | Canada | Financial, language and cultural barriers | Recruiting local community members, rural and northern community exposure, improved communication between NOSM and the local community |
| Mikhail et al [67] | Operation Remote Immunity: Exploring the impact of a service-learning elective in remote Indigenous communities | 2023 | Described how an elective in northern Indigenous communities based on service-learning influenced social accountability | Qualitative | Canada |  | Service-learning approach, personal exposure to inequities |
| Myhre et al [68] | Specialty resident perceptions of the impact of a distributed education model on practice location intentions | 2015 | Examined how the University of Calgary's distributed education training model influenced residents' practice intentions and how this differs based on speciality and program stage | Mixed methods | Canada |  | Engaging, supportive and passionate staff, cost-effective accommodations |
| Paul et al [69] | Making a difference: The early impact of an Aboriginal health | 2006 | Outlined the effects of University of Western Australia's | Cross-sectional | Australia |  | Involving local community members in curriculum |
|  | undergraduate medical curriculum |  | integration of a Aboriginal health curriculum |  |  |  | development and teaching |
| Punzalan et al [40] | The integration of primary care and public health in medical students' training based on social accountability and community-eng aged medical education | 2023 | Explored student and community perspectives regarding how socially accountable medical training incorporated primary care and public health | Qualitative | Philippines | Poor working conditions and lower salaries | Curriculum that emphasizes community immersion and hospital rotations |
| Roberts et al [41] | A longitudinal integrated placement and medical students' intentions to practise rurally | 2012 | Examined how an integrated placement affects medical students' intentions to practice in rural areas | Qualitative | Australia | Limited employment opportunities, family concerns, geographic isolation, lack of support during rural student placements | Student support during placements such as professional role models |
| Ross et al [57] | A qualitative investigation of the experiences of students and preceptors taking part in remote and rural community experiential placements during early medical training | 2019 | Described a medical program's rural community placements | Qualitative | Canada | Poor learning experiences and retention difficulties | Early remote and rural community experiential placements |
| Ross et al [58] | The training for Health Equity Network evaluation framework: A pilot study at five health professional schools | 2014 | Assessed the evaluation framework with the goal of promoting social accountability across five Training for Health Equity Network schools | Mixed methods | Australia, Canada and Philippines |  | Implementing social accountability evaluation tools |
| Sen Gupta et al [70] | Rural internships for final year students: Clinical experience, education and workforce | 2008 | Outlined the development and incorporation of a rural internship program | Qualitative | Australia | Poor quality rural placements | Well developed rural internship model to better understand local communities |
| Shannon et al [71] | Evaluation of a required statewide interdisciplinary rural health education program: Student attitudes, career intents and perceived quality | 2005 | Examined how student attitudes towards career intentions and rural health were impacted by a community-oriented rural education program | Prospective cohort | United States | Pre-existing student career intents | Community-based rural curriculum |
| Siega-Sur et al [59] | The impact of socially-accountable, community-engaged medical education on graduates in the central Philippines: Implications for the global rural medical workforce | 2017 | Explored how socially accountable professional education influenced students' career and practice intentions | Cross-sectional | Philippines |  | Tailored curriculum through community centred programs |
| Smith & Weaver [42] | Capturing medical students' idealism | 2006 | Examined the impacts of a health elective at the University of Texas Medical Branch on students | Qualitative | United States with an international elective component in Nicaragua | Declining attitudes through training, financial issues and a lack of resources | Unique mentored student experiences |
| Strasser et al [72] | Transforming health professional education through social accountability: Canada's Northern | 2013 | Outlined the effects of NOSM's social accountability mandate and described how it impacts health outcomes | Mixed methods | Canada |  | NOSM holistic curriculum involving community-partnerships |
|  | Ontario School of Medicine |  |  |  |  |  |  |
| Strasser & Cheu [73] | Needs of many: Northern Ontario School of Medicine students' experience of generalism and rural practice | 2018 | Examined student perceptions of generalism in rural practice at NOSM | Qualitative | Canada |  | Comprehensive community clerkship program |
| Tejativaddhana et al [43] | Identifying challenges and barriers in the delivery of primary healthcare at the district level: A study in one Thai province | 2013 | Described the views of stakeholders on improving primary health services and examined difficulties in care delivery in Thailand | Qualitative | Thailand | Lack of recognition of the importance of equitable healthcare access, retaining issues, economic barriers, curative rather than preventative care focus |  |
| Thompson et al [74] | Views from the field: Medical student experiences and perceptions of interprofessional learning and collaboration in rural settings | 2018 | Examined how interprofessional learning is viewed by medical students | Qualitative | Canada |  | Role flexibility, unstructured learning, positive learning environment, interprofessional learning |
| Vujcich et al [75] | “[It’s] more than just medicine”: The value and sustainability of mandatory, non-clinical, short- term rural placements in a Western Australian medical school | 2020 | Explored the rural placement program’s impact on student career intentions and community rural health needs | Qualitative | Australia | Prevailing student attitudes, | Mandatory short-term rural placements |
| Wearne et al [44] | Physical isolation with virtual support: Registrars’ learning via remote supervision | 2015 | Examined the influence of remote supervision on the educational experiences of general practitioner registrars | Qualitative | Australia and Canada | Difficulties in providing structured learning and proper supervision | Rural immersion and adequate remote supervision, giving learners a sense of responsibility |
| Wenghofer et al [45] | Impact of the rural pipeline in medical education: Practice locations of recently graduated family physicians in Ontario | 2017 | Determined how NOSM education impacted students intention to practice in rural areas and compared this to other Canadian medical institutions | Cross-section al | Canada | Lower diversity of healthcare specialties, financial limitations | Recruitment and training of rural and northern students |
| Woolley et al<br>[76] | Socially accountable medical education strengthens community health services | 2018 | Explored how those with socially accountable health professional education have influenced child and maternal health services | Cross-sectional | Philippines |  | Selective admission of representative students, service learning |
| Woolley et al<br>[77] | Does a socially-accountable curriculum transform health professional students into competent, work-ready graduates? A cross-sectional study of three medical schools across three countries | 2019 | Evaluated the impact of socially accountable medical schools on student competencies and job preparation | Cross-sectional | Austria, United States and Sudan |  | Community-based training and curriculum development |

**Table 2.** Characteristics of original articles included in the study.

| Characteristic | No. (%) |
| --- | --- |
| **Study type** |  |
| Qualitative | 24 (45.3%) |
| Cross sectional | 13 (24.6%) |
| Prospective cohort | 8 (15.1%) |
| Mixed methods | 4 (7.5%) |
| Descriptive analysis | 2 (3.7%) |
| Retrospective cohort | 1 (1.9%) |
| Retrospective case study | 1 (1.9%) |
| **Study country** |  |
| Canada | 13 (24.5%) |
| Australia | 12 (22.6%) |
| Multi-national | 7 (13.2%) |
| Philippines | 6 (11.3%) |
| United States | 5 (9.4%) |
| South Africa | 3 (5.7%) |
| Scotland | 2 (3.8%) |
| India | 1 (1.9%) |
| Japan | 1 (1.9%) |
| Kyrgyzstan | 1 (1.9%) |
| New Zealand | 1 (1.9%) |
| Thailand | 1 (1.9%) |

Studies included in this review were conducted across 15 countries: Australia, Belgium, Canada, India, Japan, Kyrgyzstan, Nepal, New Zealand, Nicaragua, Philippines, Scotland, South Africa, Sudan, Thailand and the United States. A majority of the studies were from Canada (24.5%), followed by Australia (22.6%), Philippines (11.3%), and the United States (9.4%) with muti-national studies accounting for 13.2% of the included articles. Three (5.7%) studies were conducted in South Africa (2.8%), two (3.8%) in Scotland, and one each (1.9%) in India, Japan, Kyrgyzstan, New Zealand and Thailand (Table 2).

## Thematic Analysis

### Challenges for physicians to promote social accountability

A total of 21 (39.6%) studies reported on the challenges faced by physicians promoting social accountability.^25–45^ Professional limitations and poor working conditions were identified as a barrier in nine of these 21 studies, emerging as the most prevalent challenge.^25,30,31,36–38,41,44,45^ These included long working hours^25^; higher workloads^45^; low remuneration^30,31,36,41^; absence of appropriate medical equipment and facilities to support the community and physicians’ career aspirations^30,31^; unstructured learning environment^44^; inadequate supervision^44^; and a lack of opportunities for professional development.^25,31,37,45^ Physicians who did not have a rural background were also faced with mismatches between their skills and the needs of the population they were serving, contributing to further issues within an already challenging work environment.^38^

Negative, progressively worsening attitudes among medical students and physicians were listed as a challenge in seven studies^26–28,31,32,42,43^ These were characterized by a declining sense of idealism,^26,42^ a lack of understanding and consideration for Indigenous communities,^27,28^ and diminished interest in working with underserved populations.^42^ Family medicine and primary care were poorly understood and viewed as unattractive fields that were perceived as neither valued by the public or by other physicians.^31,43^ Furthermore, physicians were reluctant to face stigma if they chose to live and work within rural communities.^32^

In addition, medical students and physicians had their own preferences of where they would like to practice as mentioned in four studies.^29,33,34,40^ The intention to emigrate was noted among individuals belonging to international, urban, and high-income backgrounds.^33,34,40^ Higher pay, better working conditions and the desire to gain experience in a foreign country were reported as the main reasons for wanting to leave.^33,40^ One finding that particularly stood out was the lack of willingness among students from very low socioeconomic backgrounds to work in underserved communities, potentially attributed to aspirations of being upwardly mobile in society.^29^

Deterrents of serving in rural and remote communities also included familial concerns, such as the scarcity of employment opportunities for partners and barriers hindering the success of their children, which were brought up in four articles.^30,36,39,41^ Moreover, geographical isolation^25,39,41^ and language barriers^25,31,39^ were cited as challenges in three studies each. A lack of amenities in rural areas, such as poor accommodation; few restaurants; and limited social activities, were mentioned in two studies^25,30^ while one study reported on the reluctance of Indigenous people to engage in projects due to prior negative experiences with medical researchers.^35^

### Challenges for medical schools to promote social accountability

Barriers to promote social accountability for medical schools and educational institutes were mentioned in 23 (43.4%) studies.^31,34,35,38–43,46–59^ Curriculum incompetency was the most frequently reported challenge and was listed in 14 articles.^31,35,39–43,50,52,55–59^ The curricula used within medical schools and clinical training were largely conventional with little focus on primary care.^40,43^ Components incorporating rural immersion were often short and lacked appropriate support mechanisms for students and physicians.^35,41^ In addition, options for medical school electives and residency tracks that incorporated community health were typically limited to individuals who were at least in their final year of medical school at which point they did not hold much utility in drawing physicians towards serving rural communities.^42^ Demands such as the need for community collaboration, tailoring the program to local needs, and the absence of an appropriate evaluation tool made it difficult to establish effective, socially accountable curricula.^35,55,58,59^ Furthermore, attempts at shifting towards a community-based approach to medical education were met with resistance from conventional medical institutions and government bodies, contributing to the current inadequacy of curricula.^52^

Financial and resource constraints to establish socially accountable medical education and training programs were discussed in eight studies.^31,34,35,42,49,51,52,55^ A lack of diversity and inclusivity in admission and recruitment policies was referred to in four studies as a barrier that limited the selection of students from rural backgrounds and those that might be more inclined to practice in these communities.^34,46,54,55^ Three studies each recognized poorly developed infrastructure^48,53,55^ and training and availability of teaching staff^31,35,55^ as a challenge for medical schools. Moreover, one study criticized the redistribution of graduate training positions for directing future primary care physicians to other specialities due to a relatively larger increase in non-primary versus primary care training.^47^

### Facilitators of social accountability for physicians

Facilitators for physicians to promote social accountability were listed in 17 (32.1%) studies.^30–38,39,44,48,54,60–63^ Community immersion, the most discussed facilitator, was recognized in 16 studies.^30,31,33–39,44,48,54,60–63^ Exposure to rural communities during their medical education (e.g., rotations) and through their own rural background and upbringing were both found to increase the willingness of physicians to serve these communities.^30,31,33,34,35,36,37,38,39,44,48,54,60,61,62,63^

Positive attitudes towards rural health and primary care were identified as facilitators in four studies.^30,31,33,60^ These included a sense of responsibility towards serving the needs of their country,^30,31,33,60^ desire to stay close to their family,^30,33,60^ attachment to their home community^33,60^ and finding the remoteness of rural areas to be appealing.^30^

The importance of peer support and establishing meaningful relationships with colleagues was highlighted in four studies.^32,36,44,60^ Similarly, the positive experience of their family members in rural areas encouraged physicians to remain and continue practicing in those communities.^36^

### Facilitators of social accountability for medical schools and medical educators

45 of the included articles (84.9%) discussed facilitators for medical schools and educators to promote social accountability.^25,27,29,30–33,35–42,45,46,49–60,62–77^ Using a community-centered curriculum _was reported in 39 studies as a successful intervention._25,27,29-33,35-37,39-42,45,49,50-52,54-57,59,60,62-64,66,67,69-77 Such curricula were designed in collaboration with community members and local physicians, specifically tailored to the needs of the area and its population.^49,59,69,76,77^ They typically entailed experiential or service learning opportunities in a rural context, including clinical rotations and _research projects._25,27,30,32,33,35,36,39,40,41,45,49,51,52,54-57,60,62-64,66,67,70-77 _Service learning activities were also_ offered with underserved, low-income and international communities.^42,56,77^ Other components of community-centered curricula included health equity,^77^ family medicine,^31^ primary care,^33,40^ public health,^40^ and interprofessional education.^50,74^

Targeted admission policies emerged as a facilitator in 11 studies.^31,38,45,46,49,53,55,60,62,65,76^ These were aimed at recruiting students that were regionally, ethnically, and socio-demographically diverse; and representative of the communities that the medical schools were serving.^62,65,76^ Strategies focused on increasing recruitment from local, rural areas; non-traditional and indigenous backgrounds; and lower socioeconomic groups, considering non-academic abilities in their admissions criteria to select students with a higher likelihood of working in primary care, remote regions and with underserved populations.^38,45,46,49,53,55,60,62,65^

The role of medical educators, acting as good role models, in supporting medical school’s social accountability mission was acknowledged in nine studies.^30,31,33,40,41,52,68,69,72^ These instructors encouraged students to consider rural practice by sharing their own passion for the field, relaying their experiences and promoting family medicine.^30,31,68,69,72^ Government support for socially accountable programs was also reported to facilitate medical schools in their implementation of these initiatives in a single study.^40^ In addition, according to the findings in one study, having an evaluative tool helped medical schools gauge their progression towards attaining social accountability, allowing them to critically analyze enabling and impeding factors and thus, work towards their goal more effectively.^58^

## Discussion

This scoping review is the first to identify social accountability practices in a rural context on a global scale. 53 publications from 15 countries documented the prevalence of rural social accountability practice, highlighting the integral significance of addressing this inequality. More understanding is needed regarding implementing social accountability in rural communities across other regions. Approximately 91% of the articles were published after the 2010 Delphi study toward a global consensus on the social accountability of medical schools (GCSA),^78^ suggesting a more significant impact set afoot.

Canadian medical schools demonstrated a heavy emphasis on social accountability practice, encompassing 24.5% of the overall literature, followed by Australia (22.6%), the Philippines (11.3%), the United States (9.4%), South Africa (5.7%), the UK (3.8%), India (1.9%), Japan (1.9%), Kyrgyzstan (1.9%), New Zealand (1.9%), and Thailand (1.9%), with multinational studies comprise 13.2%. Despite the inclusion of studies coming from diverse countries, this distribution demonstrated a lack of literature from regions of Latin America, the Middle East, Northern and Central Africa.

The practice patterns of social accountability are regionally diversified. The Christian Medical College, Vellore, Tamil Nadu, India (CMCV), has a well-documented history of practicing social accountability for over 100 years, originally started as a not-for-profit entity for services across all socioeconomic backgrounds (Jennifer Cleland, 2024). Of the included studies, 12 pertained to NOSM University.^33,34,35,39,45,57,58,62,67,72–74^ Part of NOSM’s success corresponds to its adoption of a social accountability mandate, which prioritizes the health needs of the local population and values community engagement. This mandate is guided by the belief that medical institutions bear an obligation to address the immediate health needs of the communities they serve. Key aspects of NOSM’s model include community-based learning, rural placements, Indigenous health partnerships, and a tailored student recruitment strategy.^17^

Thematic analysis indicated barriers for physicians and medical school students to contribute towards rural social accountability practice as a) inadequate working infrastructure as the most prevalent challenge, b) a mismatch between physician skills and the population’s needs, c) unfavorable attitudes of medical students and physicians, d) personal preferences attributed to socio-economic backgrounds, e) familial considerations for the development of subsequent generations due to geographical isolation, f) cultural contrasts with the local population, and g) negative inclination of Indigenous population towards medical researcher.

Barriers decelerating the spread of medical school rural practice of social accountability comprise a) inadequacies in curriculum design with insufficient consideration targeting rural settings while contenting institutional or governmental objections, b) intrinsic monetary and resource reservoir discretions within regions and communities, c) absence of admission diversity and equality considerations, d) inadequate or nascent infrastructure development, and e) human resource redistribution after graduation. Synthesized physician promotion of social accountability efforts can be summarized into the subsequent comportment: a) exposure towards rural communities, b) rural familial background, c) favorable attitudes towards rural regions during medical school studies, and d) underpinning peer-to-peer mitigation. Identified studies have focused on the implementation of medical school in mitigating social accountability challenges, encompassing a) curriculum design centering on the community and assessing the demands of the population, b) targeted admission towards recruitment of students who are inherently representative of the community or region they have the mandate to serve, c) peer, mentor, and institutional support networking through establishing role models, and d) evaluation through external tools as dimensions to measure social accountability engagement.

Evaluating the barriers and facilitators of social accountability practice has demonstrated intrinsic challenges due to the multipolar nature on a global scale, this further accentuates the demand for a unified, region-dependent, and flexible framework, extending the challenges beyond infrastructure, retention, socioeconomic status, and recruitment; encompassing philosophical landscape, and cultural differences inherent to the region.

## Limitations

Limitations of this scoping review have been identified, incorporating small sample sizes in included primary research studies, the findings reported were limited to published peer-reviewed primary studies, and all gray literature was excluded, indicating a potential overrepresentation of significant results, heterogeneity in definitions of social accountability between included studies have been considered, the inclusion criteria outlined indicated the definition social accountability in accordance to the WHO guidelines.^19^

## Conclusion

This scoping review synthesizes existing evidence from 53 articles on the barriers and facilitators of social accountability practices in rural settings across 15 countries. The identified literature captures recurring themes of medical infrastructure inadequacy, community immersion curriculum design, targeted admission, geographical isolation, and institutional or peer support. Prospective research could focus on expanding and refining current guidelines and frameworks to improve socially accountable benchmarks in rural areas.

## Statements and Declarations

### Ethical considerations

This article does not contain any studies with human or animal participants.

There are no human participants in this article and informed consent is not required.

### Consent to participate

Not applicable

### Consent for publication

Not applicable

### Declaration of conflicting interest

The author(s) declared no potential conflicts of interest with respect to the research, authorship, and/or publication of this article.

### Funding

This scoping review was conducted without financial support.

### Data Availability

The data supporting the findings of this study are available from the Supplement Material.

## Data Availability

All data produced in the present study are available upon reasonable request to the authors.

## Appendix 1: Search Strings

| Database | Query | Output |
| --- | --- | --- |
| Ovid MEDLINE | 1 exp Physician's Role/ or exp Practice Patterns, Physicians'/ or exp Physicians/ or Physician*.mp. 704701<br>2 Health Personnel*.mp. or exp Health Personnel/ 212215<br>3 healthcare provider*.mp. 38937<br>4 (doctor* or clinician*).mp. 479067<br>5 medical professional*.mp. 11818<br>6 family practice*.mp. or exp Family Practice/71016<br>7 students, medical.mp. or Students, Medical/ 45478<br>8 education, medical.mp. or exp Education, Medical/ 188730<br>9 exp Schools, Medical/ or Schools, medical.mp. 28479<br>10 primary health care.mp. or exp Primary Health Care/ 214987<br>11 1 or 2 or 3 or 4 or 5 or 6 or 7 or 8 or 9 or 10 1611375<br>12 Social Accountabilit*.mp. or exp Social Responsibility/ 25874<br>13 social responsibilit*.mp. 22707<br>14 12 or 13 27871<br>15 Rural Health.mp. or exp Rural Health/ 42036<br>16 Rural Population*.mp. or exp Rural Population/ 74618<br>17 Rural Area*.mp. 52478<br>18 Village*.mp. 36812<br>19 small town*.mp. 2091<br>20 exp Medically Underserved Area/ or underserved area*.mp. or exp Rural Health Services/ or exp Health Services Accessibility/ 154088<br>21 exp Arctic Regions/ 7600<br>22 Arctic Region*.mp. 8072<br>23 exp Health Services, Indigenous/ or exp Northern Territory/ or Northern Territor.mp. 7018<br>24 Polar region.mp. 1966<br>25 Remote area.mp. 6830<br>26 Countryside.mp. 1787<br>27 Rural.mp. or exp Hospitals, Rural/ 212740<br>28 Remote*.mp. 121675<br>29 15 or 16 or 17 or 18 or 19 or 20 or 21 or 22 or 23 or 24 or 25 or 26 or 27 or 28 490434<br>30 11 and 14 and 29 841<br>31 limit 30 to yr="2000 -Current" 475 | 475 |
| Scopus | TITLE-ABS-KEY ( ( "Physician*" OR "Health Personnel*" OR "Medical Doctor*" OR "Healthcare provider*" OR "Clinician*" OR "Medical practice*" OR "Medical student*" OR "Medical Education*" OR "Healthcare*" OR "Primary Health Care*" OR "Medical professional*" ) AND ( "Social Accountabilit*" OR "Social Responsibilit*" ) AND ( "Rural health*" OR "Rural Population*" OR "Rural Area*" OR "Remote | 245 |
|  | Area*" OR "Arctic region*" OR "Northern Territory*" OR "Rural Health Service*" OR "Indigenous Health" OR "remote" OR "rural" OR "Medically underserved area" OR "Indigenous Health Service*" OR "Countryside") ) AND PUBYEAR > 1999 AND PUBYEAR < 2025 |  |
| CINAHL | (("Physician*" OR "Health Personnel*" OR "Medical Doctor*" OR "Healthcare provider*" OR "Clinician*" OR "Medical practice*" OR "Medical student*" OR "Medical Education*" OR "Healthcare*" OR "Primary Health Care*" OR "Medical professional*" ) AND ("Social Accountabilit*" OR "Social Responsibilit*" ) AND ("Rural health*" OR "Rural Population*" OR "Rural Area*" OR "Remote Area*" OR "Northern region*" OR "Arctic region*" OR "Northern Territor*" OR "Northern health*" OR "North America*" OR "Northern communit*" OR "Rural Health Service*" OR "Indigenous Health" OR "north" OR "Remote" OR "arctic")) | 445 |
| Web of Science | ((("Physician*" OR "Health Personnel*" OR "Medical Doctor*" OR "Healthcare provider*" OR "Clinician*" OR "Medical practice*" OR "Medical student*" OR "Medical Education*" OR "Healthcare*" OR "Primary Health Care*" OR "Medical professional*") AND ("Social Accountabilit*" OR "Social Responsibilit*") AND ("Rural health*" OR "Rural Population*" OR "Rural Area*" OR "Remote Area*" OR "Northern region*" OR "Arctic region*" OR "Northern Territory*" OR "Northern health*" OR "North America*" OR "Northern communit*" OR "Rural Health Service*" OR "Indigenous Health" OR "northern" OR "remote" OR "rural")) | 149 |
| Ovid EmBase | ((("Physician*" or "Health Personnel*" or "Medical Doctor*" or "Healthcare provider*" or "Clinician*" or "Medical practice*" or "Medical student*" or "Medical Education*" or "Healthcare*" or "Primary Health Care*" or "Medical professional*") AND ("Social Accountabilit*" or "Social Responsibilit*") AND ("Rural health*" or "Rural Population*" or "Rural Area*" or "Remote Area*" or "Northern region*" or "Arctic region*" or "Northern Territor*" or "Northern health*" or "North America*" or "Northern communit*" or "Rural Health Service*" or "Indigenous Health" or "north" or "Remote" or "arctic")).mp. [mp=title, abstract, heading word, drug trade name, original title, device manufacturer, drug manufacturer, device trade name, keyword heading word, floating subheading word, candidate term word] | 121 |

## Appendix 2: Inclusion & Exclusion Criteria

|  |  |
| --- | --- |
| **Participants:**<br>Studies with or without human participants of any demographic that address social accountability among physicians/medical schools.<br>- Studies should either be discussed from a healthcare perspective by physicians or medical school faculty.<br>- We will include studies that make generalized claims in these two areas without directly mentioning human participants. | Exclude studies that don't discuss social accountability from the perspective of physicians or medical schools/medical education.<br>Studies not involving physicians or not relevant to medical education. |
| **Setting:**<br>Studies that address the issue of social accountability within a rural/remote community | Challenges/facilitators not listed.<br>Exclude studies that talk about promoting social accountability in any other settings (e.g. densely populated cities, non-rural) |
| **Study Design**<br>Primary articles of any study design that addressed social accountability among physicians and medical schools in the context of rural and remote communities. | Non-research articles (e.g., errata, commentaries, book chapters, letters to the editor, protocols, editorials, perspectives and opinion pieces), thesis dissertations, conference proceedings, and reviews. |
| **Languages:** English | Exclude papers in other languages |

## Appendix 3: Data Extraction Form

|  |
| --- |
| Title of study<br>Lead Author<br>Print Publication Date |
| Aim of study (objectives)<br>Study Design (e.g. qualitative, mixed-methods)<br>Country of study (e.g. Canada, US)<br>Setting (e.g. rural village, medical institution) |
| Population/group studied (e.g. medical educators, physicians, medical students)<br>Population size (if applicable)<br>Population demographics (e.g. female physicians)<br>Population age |
| How does the study define social accountability?<br>How does the article describe the importance of social accountability?<br>(Do they discuss how social accountability impacts patient outcomes or community well-being?) |
| How does the study measure social accountability (e.g. interviews/perspectives, surveys, other metrics)<br>Challenges listed for physicians to promote social accountability<br>Challenges listed for medical schools/medical education to promote social accountability<br>How do these barriers affect social accountability? |
| Are there any proposed solutions to social accountability challenges?<br>Does the author mention any key roles that physicians should play?<br>Does the author mention any key roles medical schools or medical educators should play?<br>Are there any facilitators that are discussed? (e.g. successful programs)<br>Does the intervention have an impact on social accountability?<br>Does it mention how the successful implementation of social accountability can be mirrored in other communities? (e.g. similar medical education training frameworks, physician workshops - how can these facilitators be translated to other communities)<br>Does the author make any other recommendations? |
| Article Conclusions<br>Study conclusions made<br>Does the article suggest any future research directions?<br>Study limitations<br>Other interesting information |

## Notes

### Competing Interest Statement

The authors have declared no competing interest.

### Funding Statement

This study did not receive any funding.

